# Assessing the epidemic impact of protests during the COVID-19 pandemic

**DOI:** 10.1101/2022.10.10.22280896

**Authors:** Inho Hong, Leonardo N. Ferreira, Alex Rutherford, Manuel Cebrian

## Abstract

Protests during the COVID-19 pandemic present a complex trade-off between democratic rights of freedom of assembly and an epidemic risk, and have created a need for careful assessment of protest-driven infections. Here, we build a coupled disease transmission model and assess the impact of protests on the COVID-19 spread in the continental US using a dataset of 4,121 protests and 1.66 million protesters between April and June of 2020. We find that protests in 2020 had limited effects, creating tens of additional daily cases country-wide, due to their small size. However, a simple scaling relation of protest-driven infections derived from our simulations reveals that very large protests with over millions of participants can significantly boost outbreaks and impact the healthcare system. In the worst-case scenario, very large protests can add over 20,000 daily cases and over 7,000 ICU admissions over the continental US. We hope our model can aid the policy rationale to maintain freedom of assembly in the current and future pandemics, while providing estimates for preparations for a healthcare surge in the worst-case setting.

## Introduction

Since the beginning of the COVID-19 pandemic, a large number of protests have happened over the world for many different reasons. Some of the protests were directly motivated by the COVID-19 impacts and policies such as economic downturn, closure of businesses, mobility restrictions, social distancing and lockdown^1–5^, while many other protests were related to long-standing social issues such as racism, labor, or politics^6^, represented by the George Floyd protests on a large scale. As seen in these examples, the emergence of protests is a very complex phenomenon affected by many different factors including online ecology^7^, momentum^8^, contagion^9^ and protesters’ attributes^10^, and thus protests have a highly dynamic nature^11–13^ and a high variance in the size described by a power-law distribution^14^.

Protests during a pandemic have a complex dilemma that large gatherings may involve a potential risk of epidemic spreading while protesting is an important democratic right^6^. The uncertainty of the epidemic risk led to debates over restricting protests for disease mitigation and guaranteeing freedom of expression^15–17^. Additionally, protests directly related to COVID-19 policies involve a potential risk by a feedback loop of epidemic-driven protests and protest-driven epidemics. While this protest-driven epidemic spreading has been anticipated, its assessment has been obstructed by the dynamic nature of protests, identification and contact-tracing of protesters, and the lack of a model tuned for disease transmission at protests. As such, the statistical relation between protests and an increase in patients is not conclusive^18–21^. If a model can assess the scale of new infections and the resulting impact on the hospital capacity for a given protest size, it could help policy makers find a balance between pandemic mitigation and the democratic right to protest, and be prepared for an upcoming increase of patients and mitigation strategies such as devoting more resources to special contact-tracing^22^ for protest-like events.

Compartmental disease transmission models, describing epidemic dynamics with a set of simple differential equations, can be used to estimate the number of infected individuals in such large gatherings^23^. Compartmental models have been widely applied for estimating those epidemic risks and extensively studied with increasing availability of mobility networks derived from empirical data and the emergence of COVID-19^23–28^. While the compartmental models have provided reliable projections on the epidemic trajectory, we still lack a model to estimate how protests may affect the overall epidemic trend on a national scale.

In this article, we build a coupled compartmental disease model optimized for protest-driven epidemic spreading and estimate the impacts of protests on the epidemic trend and hospitalization. Specifically, we focus on protests over COVID-19 in the US to examine the impact of protests solely driven by the pandemic, as well as the impact of all protests. Then, we estimate the impacts on new infections and hospitalization by different scales of protests and epidemic situations. By doing so, we uncover 1) the limited impacts of protests during 2020, 2) the critical protest size in the order of millions in a total population of the US to make significant changes in the epidemic trend, 3) a simple scaling relation between protest-driven infections, protest size, and epidemic size, and 4) policy implications on next pandemic waves, a feedback loop of protests and epidemics, and extension to the other types of outdoor gatherings.

### Modeling protest-driven epidemics

Here we describe the coupled disease transmission model to estimate the impacts of protests on epidemic spreading. Overall, this model divides a population into two sub-populations of protesters and non-protesters at the level of counties (second level administrative divisions in the United States), calculates compartmental disease spreading separately for each group, and aggregates the updates for each day (see Fig. 1a). For non-protesters, we adopted the SEIR disease transmission model with mobility, well known as the Global Epidemic and Mobility Model (GLEaM)^24^ to incorporate spatial disease spreading, and modified it for domestic mobility including commuter flows and airline traffic. For protesters, we assumed that protesters in each county of a state gather at a unique site of the state, go for protests, and return to their home counties. Accordingly, we used a simple SEIR model without mobility for protesters. The model gives the same results for a single large protest and many small protests in a state for the same total number of protesters. The number of daily protesters at each county was estimated from the number of daily protesters at the state level based on the protest data from Count Love^30^ which was intensively reviewed by Fisher et al^10^. The initial size of each compartmental population was estimated using the county-level COVID-19 statistics^29^ (See Methods and SI. Section S2 for more details).

**Figure 1.**
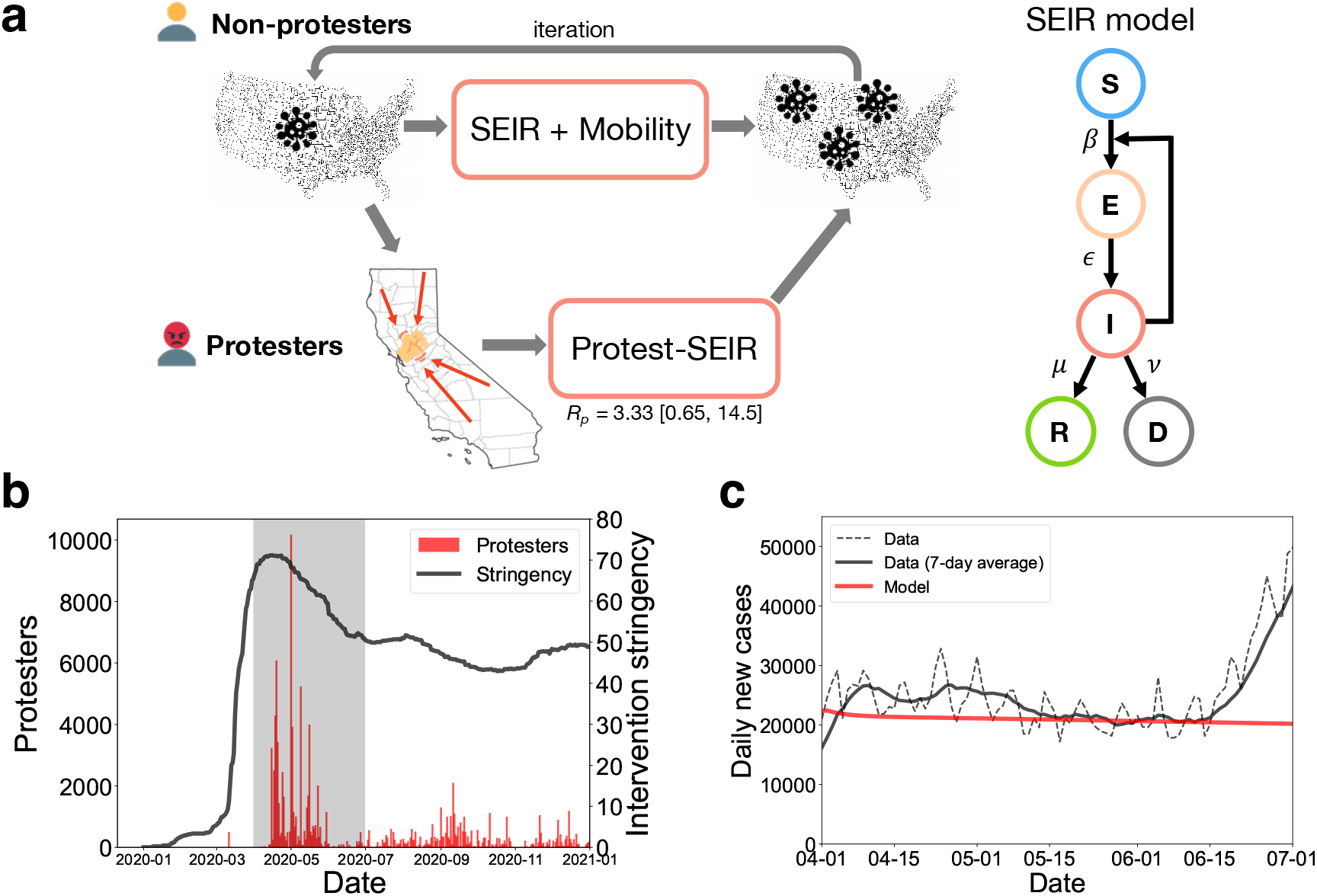
The model design and the trends of protests, interventions and COVID patients. **a**. The schematic of the coupled disease transmission model. (Left) The epidemic spreading among non-protesters and protesters are calculated separately using the SEIR model with mobility and the ordinary SEIR model, respectively, for each day. We assumed that protesters from each county of a state gather at a site for protesting. The number of protesters at the county-level is estimated as the product of the state-level protesters and the fraction of county population. The updated compartmental population is summed up for each day. (Right) The SEIR model describes the epidemic dynamics between the compartments of susceptible, exposed, infected, recovered and dead population, represented by the first letter of each compartment (Details in Sec. Model). **b**. The daily total number of protest attendees for protests over COVID-19 policies in the US (red) and intervention stringency (black). Intervention stringency stands for the number of different mitigation measures. The gray shade indicates the period when the protests were most active. **c**. The baseline epidemic estimation for non-protesters. The black curves show the total daily new confirmed cases in the US between April 1 and July 1 of 2020 with fluctuations corresponding to weekends^29^ (dotted for raw data and solid for 7-day averages). The red curve shows the estimation by our epidemic model for the reproduction number of 1.05.

One key factor to distinguish the dynamics of disease spreading in protesters and non-protesters is the different transmission rates attributed to different behavioral and epidemiological properties of protests. The transmission rate among non-protesters can be estimated from the reproduction number of the epidemic trend, however, estimation of the transmission rate of the coronavirus among protesters requires some assumptions since we lack reliable empirical measures under protest conditions. We estimated the reproduction number at protests based on two unique features: 1) low outdoor transmission and 2) close contacts in high-density crowds. The outdoor transmission rate of COVID-19 is still uncertain, however several early studies reported that indoor transmission is much higher than outdoors with 18.7 times higher rate and the confidence interval of 6 to 58 times^31^. This low outdoor transmission rate may be considered as a reason for epidemiological safety of protesting, however, close contacts should also be considered for this evaluation. Since the 2-meter social distancing rule is generally used despite debates on its effectiveness^32^, we estimated the number of people within a radius of 2 meters in a high-density crowd of protesters using Jacob’s crowd formula^33^ which is widely adopted in estimating a crowd density of protests^34,35^, consistent with observations from protests^36,37^ and large-scale events^38^. As a result, the number of protesters with close contacts may vary from 12.5 to 29 with the central value of 20.75. Therefore, by combining the low outdoor transmission rate, the size of crowds, and the base reproduction number of COVID-19 without interventions, i.e., *R*_0_ = 3.0^39^, we get the estimated reproduction number at protests as 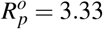 with the lower and upper bounds of 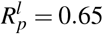 and 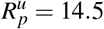, respectively. This estimated range is consistent with observations in high-risk superspreading events^40–42^.

Using a dataset of protest events from Count Love^10,30^ and intervention stringency from OxCGRT^2,43^, we characterize the temporal patterns of protests over COVID-19 policies in the United States in 2020 to see how protests evolved in response to COVID-19. Figure 1b shows the trends of the total number of protesters and intervention stringency in 2020. After intervention stringency reached its peak on April 14 after the sharp rise in March, protests over COVID-19 policies started to emerge and reached the peak of approximately 10,000 protesters on May 1. At the state level, the maximum protest occurred 24 days later than the date of the maximum intervention stringency on average. The protests were most active during the period from April to June, and accordingly, our analysis in this paper focused on the dynamics in this time window from April 1 to June 30 of 2020. During this period, 68,670 cumulative protesters participated in 389 protests over COVID-19 policies, and there were 1,661,215 protesters in 4,121 protests for all types of protests. The number of protesters includes double counting since each individual protester cannot be identified in the data.

To estimate protest-driven infections, the baseline epidemic dynamics among the general public should be identified. Specifically, one should get the reproduction number that best describes the epidemic trend in the 3-months period from April 1 to determine the transmission rate *β* for the model. In this period, the number of daily new cases was approximately 20,000, and the trend was relatively stable in time. Since the causal relation between protests and COVID incidence is not clear^18–21^, we estimated the baseline reproduction number using the epidemic trend, assuming that it represents the dynamics among non-protesters. As a result, the reproduction number of 1.05 best described the baseline epidemic trend for non-protesters as shown in Fig. 1c (See Supplementary Fig. S1 for the model trend for different reproduction numbers).

### Limited impact of protests on epidemics

Using our model and the dataset of protests and epidemic incidence in the US, we estimate the spatial and temporal excess incidence of COVID-19 patients by protests over interventions during the period from April 1 to June 30. We estimated this excess incidence by subtracting the estimation without protests from the estimation with protests by regarding the simulation without protests as a null model. Therefore, the excess cases include indirect infections in non-protesters from infectious protesters in their home counties.

Figure 2a shows the estimated protest-driven infections by protests over COVID-19 policies. The number of cumulative cases during the 3-months period is in the order of 100 at a maximum. This number, corresponding to a few excess cases per day, is very small compared to the total daily incidence of around 20,000 cases. This effect is also small compared to other interventions. For example, the effect of stay-at-home orders was estimated as a 30% reduction in incident cases^44^. The map in Fig. 2a presents the cumulative number of protest-driven infections at the state level. Some states where protests over COVID-19 policies were more active or lasted longer show higher incidence than the other states; for instance, Michigan is the state where anti-lockdown protests started, and California is the state that had the largest anti-lockdown protest. Although we observe some regional differences by different patterns of protests at the state level, the impact on the epidemic trend was little compared to the whole epidemic trend.

**Figure 2.**
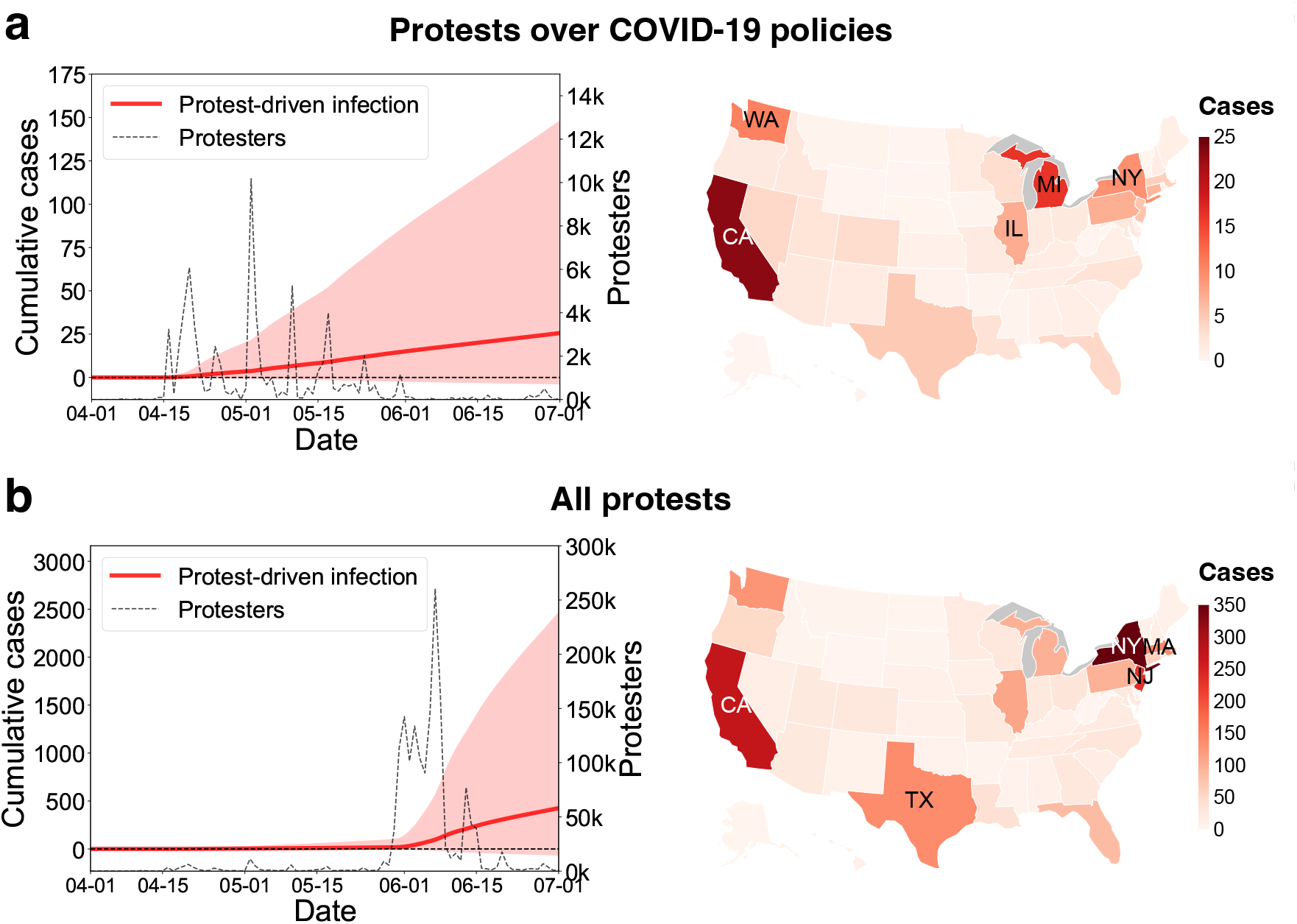
Estimated protest-driven epidemics. **a**. Estimated excess infections by protests over COVID-19 policies: the nationwide daily trend (left) and the state-level incidence (right). In the trend, the black dotted curve and the red curve depict the time series of protesters over COVID-19 policies and the cumulative number of the excess protest-driven infected cases estimated by the model, respectively. The red line denotes for the reference reproduction number (i.e., 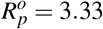), and the red shade denotes its deviations for the upper (i.e., 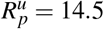) and lower (i.e., 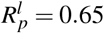) limits of the reproduction number. The color on the map denotes the cumulative of estimated excess infections in each state on June 30, 2020. The top 5 states with highest protest-driven incidence are indicated on the map. **b**. Estimated excess infections for all protests: the nationwide daily trend (left) and the state-level incidence (right). The top 5 states with highest protest-driven incidence are indicated on the map.

This limited impact of protests over COVID-19 policies is consistent with apparently no significant increase after a series of the protests. The reasons for this small impact are the low number of infected participants in the protests and the relatively small size of the protests compared to the total population. Our model estimates that there were 13 infected individuals in 10,000 protesters on average, and this number is small compared to the whole population of patients. Therefore, our findings confirm that the little impact of the protests on disease spreading was attributed to its small size.

The results from protests over COVID-19 policies lead us to a question - what if the protest were larger? To examine this, we simulated the spread of COVID-19 in all types of protests within the same time window from April 1 to June 30 of 2020. In this period, there were large protests dominated by George Floyd protests. A simulation of disease spreading in such large-scale protests can answer how large protests may impact the overall epidemic trend.

Figure 2b depicts the number of protesters and the cumulative excess cases by all protests. The number of daily protesters reached around 300,000, and accordingly, the excess cumulative cases were in the order of thousands after one month from the surge of protesters. The cumulative cases corresponds to daily excess incidence of 13.4 cases for the base reproduction number (i.e., 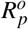), and 77.5 cases for the upper limit (i.e., 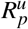) during June 2020. This number is marginally comparable to overall daily incidence, ∼20,000 cases per day. A state-wise comparison in Fig. 2b shows similar tendency with the result for protests over COVID-19 policies, while the cumulative excess cases are in the order of hundreds but still insignificant at the state level.

From our simulation, large-scale protests in 2020 had limited impacts on the epidemic trend due to the low outdoor transmission rate despite the large protest size. Although the excess daily infected cases around one hundred is not negligible in the aspect of public health, this is still small to drive a new surge compared to the total number of daily incidence. It appears that this intermediate level of excess cases by protests led to a inconclusive statistical significance on protest-driven disease spreading^18–20^.

### Critical protest size for protest-driven infections

Our simulation based on the protest data indicates that protests in 2020 had limited impacts on the epidemic trend due to its small size. Then, what is the critical size of protests that makes a significant epidemic impact? To answer this question, we multiplied the number of protesters over COVID-19 policies, and examined the protest-driven excess cases for different levels of the multiplication factor. Since we aimed to investigate the scaling effect by protest size, temporal patterns of the multiplied trend were set to be identical with the original trend (see Fig. 3a for an example of multiplication by a factor of 3).

**Figure 3.**
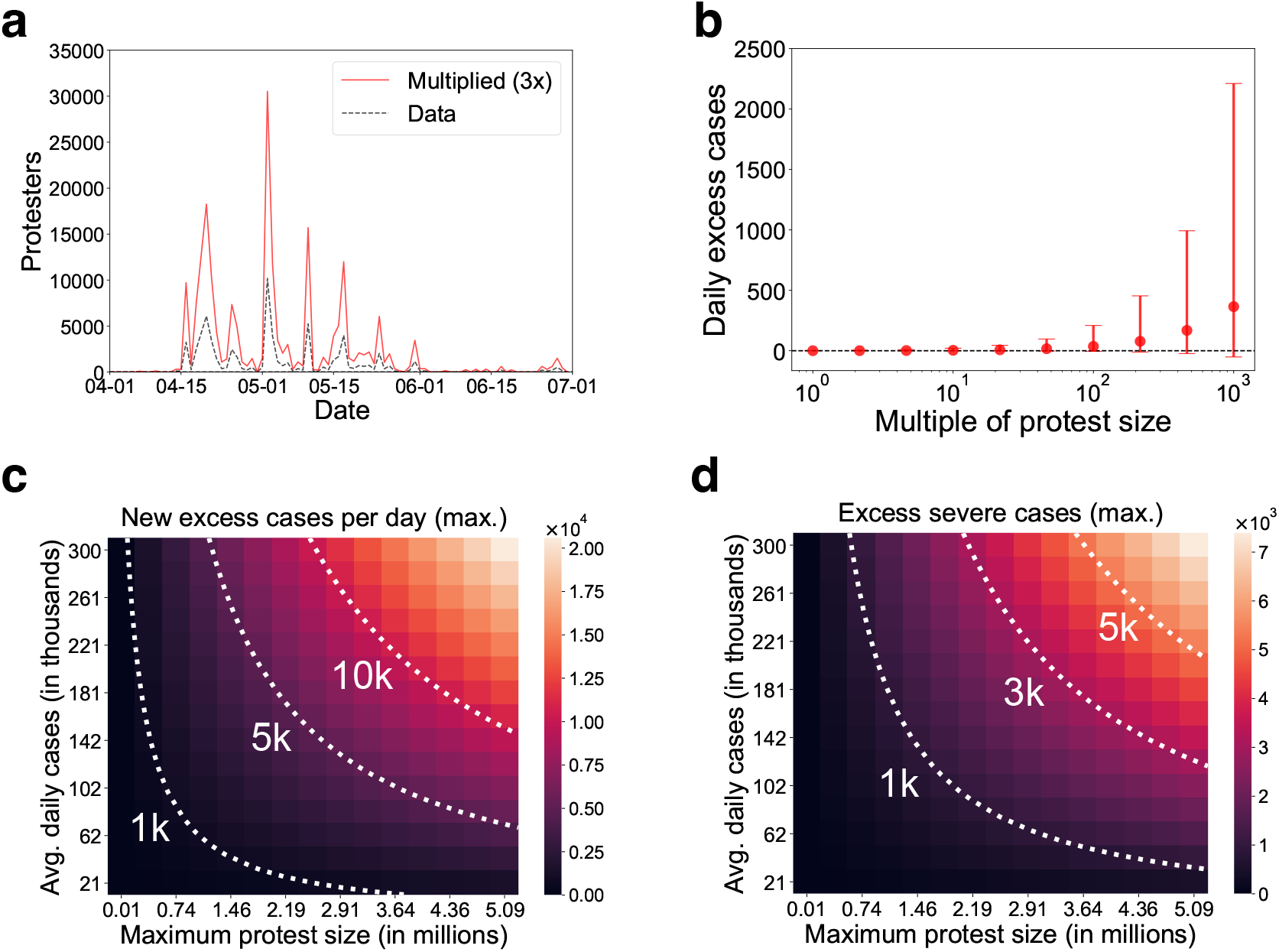
Excess cases for various scales of protests and epidemics. **a**. Multiplication of the protest size. We multiplied the number of protesters of each state by a multiplication factor (e.g., 3 in this figure) on the trend of protests over COVID-19 policies. **b**. The estimated excess daily cases on June 30, 2020 by different multiplication factors of protests. The error bars denote the simulation results for the upper and lower limits of the reproduction number. **c-d**. The excess infected cases (c) and the excess severe cases (d) for different levels of protests and epidemics. The values stand for the maximum of daily estimation for the upper limit of the reproduction number.

As a result, Fig. 3b shows monotonically increasing excess cases with increasing protest size. As a monotonically increasing trend does not have a specific critical point, we regarded the critical protest size as the point where the size of excess cases takes a percent of the total daily cases to make a significant impact. This critical protest size is found at the multiplication factor in the order of 100 corresponding to 1 million maximum daily protesters in total across the entire country. More precisely, the critical size is located at around 1 million for 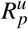, and also located at 5 million for 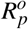 for the entire US. This critical size is large but still feasible as observed in large-scale protests and mobilization events in the last decade. For example, Women’s March in 2017, likely the largest single-day protest in US history, was estimated to have recruited approximately 3 million to 5 million over the entire US^45^. Also, Women’s March^46^ in 2018, March for Our Lives^47^ in 2018, and the George Floyd protests^30^ in 2020 were estimated to have recruited around 2 million, 1.2 million, and 0.3 million on a single day, respectively. This estimation of the critical protest size suggests that protests in 2020 were not large enough to make a significant change in the epidemic trend since its size was smaller than the critical size in the order of millions, despite its large scale.

Since our estimation is based on a meta-population model, the excess cases driven by protests are necessarily depending on the proportions of protesters and infectious people in the total population. Our model indicates that the scale of excess infections and following additional pressure on the hospital capacity can be threatening while the proportion of excess cases remains at a small proportion. For this understanding, we estimate the scale of protest-driven excess infections and the following excess severe cases by multiplying both of the protest trend and the epidemic trend. The simulation covers up to 15 times multiplication, equivalent to 300 thousand daily cases, for the epidemic trend since it was the maximum level of incidence before the spread of the Omicron variant^48^.

The estimation of daily excess cases by different levels of protests and new incidence in Fig. 3c shows that large protests at the epidemic peak can add daily patients in the order of ten thousands. Also, even smaller protests with 100 thousand protesters turned out to add a thousand new patients per day. This result implies that the small scale of infections at protests in 2020 was attributed to the epidemic situation with relatively lower incidence, and it could have been more severe if large-scale protests occurred at the peak of COVID-19.

This multiplicative nature of protest-driven infections can be formulated as a rule-of-thumb scaling relation of protest and epidemic sizes. Precisely, protest-driven excess cases Δ*I*_*p*_ can be expressed as a function of the total number of protesters *P*, the total population *N* and the total daily incidence Δ*I*_*t*_ according to:

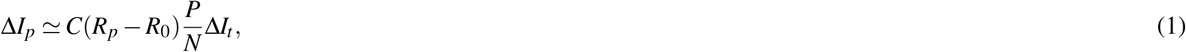

where *R*_*p*_ and *R*_0_ are the reproduction numbers for protesters and non-protesters, respectively (see SI. Section S3.2 for validation). The constant *C* is calculated as 0.331 for the maximal cases depicted in Fig. 3. According to this relation, the critical protest size *P*_*c*_ can be derived as

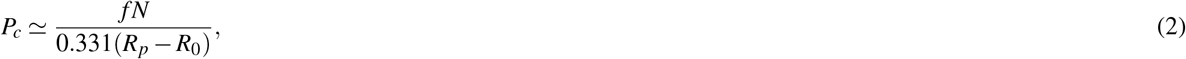

where *f* denotes the critical fraction Δ*I*_*p*_*/*Δ*I*_*t*_ = 0.01. For the base reproduction number for protests 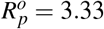, the critical protest size is given as *P*_*c*_ = 4.39 10^6^, and for the upper limit 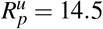, the critical size is *P*_*c*_ = 7.45 10^5^, confirming the estimation from Fig. 3b. Thus, we suggest that the 0.7 million to 4.4 million protesters on a single day over the entire US is the protest size that can create a significant epidemic impact given the reproduction number of COVID-19 and the total US population, 331 million. Furthermore, this scaling relation can inform the critical protest size as a rule-of-thumb estimate for other countries with different scales of population. For instance, in the case of Germany with a population of 83 million, the critical protest size is estimated as 0.3 million to 1.7 million, supposing that all the other conditions are fixed. In addition, the scaling relation shows that a small variation in the reproduction number can lead to a completely different scenario of disease spreading; for example, a small increase in 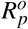 and 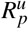 can drastically decrease the critical size of the critical protest (see SI. Section S3.3 for the varying critical size by different levels of *R*_*p*_).

Based on this estimation of excess cases, we examine the potential impacts on the hospital capacity by protests at different epidemic situations. For simplicity, we assumed that 2.5% of active infectious cases are subject to severe cases who need hospitalization in intensive care units (ICU) according to the statistics on the first COVID-19 wave^49^, and we counted the maximum number of excess active severe cases added by protests. Figure 3d shows that the excess severe cases may range from a thousand to 7 thousand if protests occur at an epidemic peak. This number takes up a significantly large fraction around 20% of the total ICU capacity which recorded maximum 30 thousand beds for COVID patients in the US^50^. Therefore, mass protests at an epidemic peak can burden the ICU capacity by a sharp increase of severe cases.

## Discussion

While epidemic spreading in protests during the pandemic has been in debates over the democratic right and epidemic containment, not many features were known about the substantive risks of protest-driven epidemics. This study provides a model for estimating the impact of protests on the overall epidemic trend by modifying the SEIR disease transmission model. As a result, we find that protests in the US in 2020 created limited impacts on epidemic spreading, and that a significant epidemiological impact can be made when the daily protesters are in the order of millions. We also inform that, in the worst-case scenario when large protests happen at the epidemic peak, protests may increase severe cases in thousands and lead to a critical burden to the healthcare capacity. Therefore, these findings highlight the need for an adaptive containment strategy for protests in response to the present scale of a disease outbreak.

Our findings inform the existing risks of epidemic outbreaks by mass protests in the near future. On the one hand, there were many mass protests larger than the critical size, i.e., millions of protesters, in history^45–47^, and the power-law distribution of protests implies a possibility of even larger protests^14^. On the other hand, large-scale pandemics such as COVID-19 might be recurring in the near future^51^. Our model can also be useful for those large protests in future pandemics.

While this study focused on the epidemic impacts of street protests, application of this model can be extended to any large outdoor events and even to indoor events by adjusting the crowd density, the crowd size, and the transmission rate. This application may include festivals, sport games, concerts and elections^22^. The estimation is expected to contribute to establishing appropriate preventive measures for those events, such as limiting the density of crowds as well as the maximum size. For example, anonymous contact tracing apps can be useful to inform the increased risk after a large gathering by sideward contact tracing^52^ that detects random encounters in the same group.

A feedback loop of protests and epidemics is another important feature that should be considered in making containment policies over a disease. Many protests such as anti-lockdown protests were a direct consequence of containment measures. If those protests grow to a large scale, they may drive another epidemic waves, and in turn, intensify restrictions, induce other protests, and recursively exacerbate the pandemic. This feedback loop is more feasible in a society under higher social unrest where containment measures are more likely to induce mass protests. Therefore, policy makers should carefully consider the increased level of social unrest since the pandemic when they decide stringency of containment policies. Although the substantive risk of this feedback loop remains unexplained, its quantitative assessment is still challenging. A proper estimation of the epidemic impact by the feedback loop is only possible with a reliable quantitative model on how intervention policies cause protests at a specific scale, timing, and location. The emergence of protests by intervention policies can be influenced by multiple factors such as individuals’ perception on interventions^53^, actual socio-economic losses^5^, and offline and online mobilization^7,54^, and thus it requires following studies in the aspects of behavioral science, political science and data science. We expect the study of the feedback loop and the protest evolution as an exciting future direction.

We emphasize that this paper suggests a necessary condition for epidemiological safety, not a sufficient condition. This study focused on the quantitative features of protests such as the size, density, and transmissibility, but some potential risks by behavioral factors could not be included in the model. For example, protesters may have pre-protest indoor gatherings that might lead to incubation of the disease and another epidemic wave^55^. Therefore, the estimation from our model should be interpreted as a condition for informed risk, not for guaranteed safety. The critical size of millions of protesters for the US indicates a definitive risk of protests larger than this point, but does not mean epidemiological safety of protests smaller than the critical size.

This paper is not without limitations. Since the home location of each protester and the exact locations of protests are not available from data, we assumed that protesters in a state gather at a site, which may lead to overestimation of protester mobility. Also, the number of protesters in each county was assumed to be proportional to the county population, meaning that the heterogeneity in tendency to protest across different counties in a state was not incorporated in the model. Next, behavioral risks in disease spreading were not included in the model. Since our model is a meta-population model, it is hard to model some protest-related behaviors that may affect disease spreading such as indoor gatherings before protesting, long-distance traveling to and from protests, and accordance with hygiene rules during protests. Nevertheless, since those behavioral patterns are likely to be net risks rather than benefits in terms of epidemiological safety, our model is still useful for anticipating the minimal epidemic impacts of protests. Lastly, we limited our model to the simplest form without including age groups, vaccination, quarantine and hospitalization. Inclusion of these factors will allow better estimation for more specific situations.

## Methods

### Data

The protest-driven epidemic model in this paper includes a few different types of data: state-level protest trends, mobility flows, intervention stringency, epidemic trends, and demographics.

The state-level protest data was obtained from Count Love^30^ for the period of 2020. It includes each protest’s date, number of attendees, location, and tags. Due to standardization, we aggregated the data to the state level. We identified the COVID-related protests by tag “Coronavirus”, and further found protests over COVID-19 policies for those which include one of the following tags, “Against pandemic intervention”, “Against closure/relocation”, or “Against vaccines”. For more details, see SI. Section S1.

To incorporate both short- and long-distance travels in the epidemic model, we combined the US county-wise commuting data and the US airport-to-airport aviation data. The county-wise commuting data was obtained from 2011-2015 5-Year ACS Commuting Flows of US Census (https://www.census.gov/data/tables/2015/demo/metro-micro/commuting-flows-2015.html). The domestic aviation data was obtained from the US Bureau of Transportation Statistics for the travels in 2019 (https://transtats.bts.gov). As the aviation data is based on the number of travelers between airports, transformation to county-wise flows is required to combine it with the commuting data. We estimated the county-wise airline traffic using the gravity model on the traffic volume and the distance. The summation of aviation mobility and commuting is used as the county-wise mobility flows. Also, we assume that the mobility flow in our scope is 60% of the pre-COVID mobility^56^.

We obtained the intervention stringency index data for US states from the Oxford Covid-19 Government Response Tracker (OxCGRT) (https://github.com/OxCGRT/USA-covid-policy)^43^. This stringency index is based on the number of different countermeasures in effect. In a few different definitions, we used the index “StringencyIndexForDisplay” in which daily variations were removed by moving average.

The COVID-19 statistics in the United States based on reports from state and local health agencies was downloaded from The New York Times (https://github.com/nytimes/covid-19-data)^29^. The data contains the number of cumulative confirmed cases by counties. The demographic information of counties was obtained from the US Census Bureau (https://data.census.gov/cedsci/). Our simulation is based on the total 3141 counties in 50 states excluding territories and two counties without commuting data.

### Model overview

The overall flow of the coupled disease transmission model in this paper includes: 1) dividing population into protester and non-protester groups, 2) simulating the epidemic dynamics for non-protesters and protesters separately, and 3) aggregating the results. The disease spreading model includes 7 different compartments, i.e., susceptible (*S*), exposed (*E*), asymptomatic infectious (*I*^*a*^), traveling symptomatic infectious (*I*^*t*^), non-traveling symptomatic infectious (*I*^*nt*^), recovered (*R*), and dead (*D*) for each county and each (non-)protester group. We set the proportion of asymptomatic population *p*_*a*_ = 0.2, the proportion of traveling population in symptomatic population *p*_*t*_ = 0.5, and the transmission reduction factor of asymptomatic individuals *r*_*β*_ = 0.5^24,57^.

### Dividing protesters and non-protesters

The number of protesters *P*_*i*_(*t*) in county *i* on day *t* was assumed to be proportional to county population *N*_*i*_(*t*) and state-level protester size *P*_*s*_(*t*) given by data as

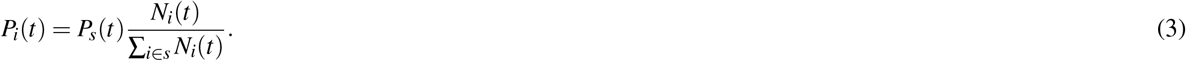

Accordingly, the non-protester size *Q*_*i*_(*t*) is given as *Q*_*i*_(*t*) = *N*_*i*_(*t*) − *P*_*i*_(*t*). Each epidemic compartment size is also proportionally assigned for 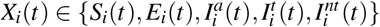 as

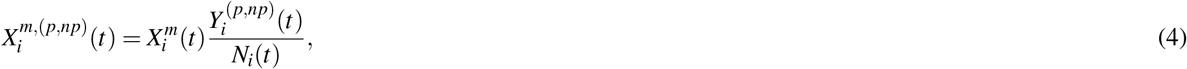

where 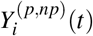 denotes the size *P*_*i*_(*t*) or *Q*_*i*_(*t*) by the choice of protesters (*p*) or non-protesters (*np*).

### Baseline epidemic model for non-protesters

For the epidemic simulation for non-protester groups, we adopted the SEIR compartmental disease spreading model with mobility^24^. While this model is based on the simple SEIR model, the model has a modification in the transition rate *λ*_*i*_ from the susceptible to the exposed by including mobility from the residential area and the returning rate. For county-wise daily travel matrix of commuting 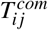 and airline travels 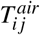 from county *i* to county *j*, the visiting rate *σ*_*i j*_ and the effective number of visitors *v*_*i j*_ are given as

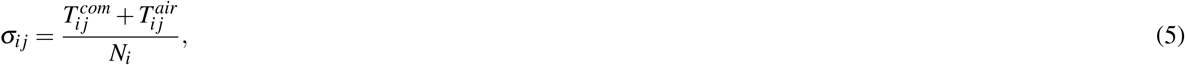

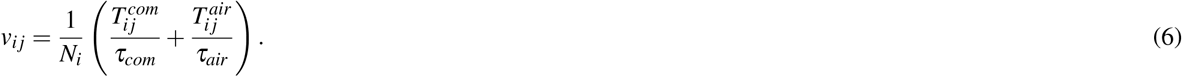

We assume different returning rates of commuters and airline travelers as *τ*_*com*_ = 3 day^−1^ and *τ*_*air*_ = 1/14 day^−1^. The summation of *v*_*i j*_ over all origins is denoted by *v* _*j*_ = ∑_*i*_*v*_*ij*_. Then, the transmission rate *λ*_*j*_ in county *j* given in the reference^24^ can be simplified as

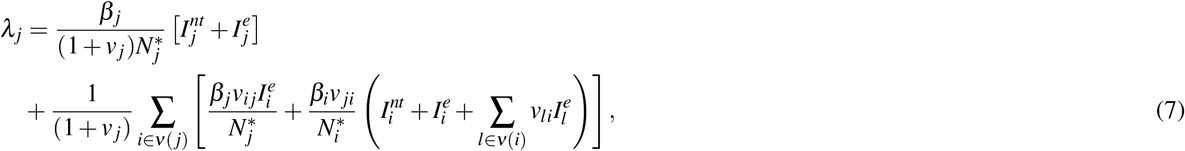

where 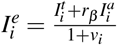 and 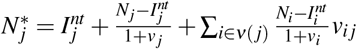

We assume that the transmission rate without mobility *β*_*j*_ is uniform across different counties. Then, the transmission rate can be estimated from the reproduction number *R*_0_ as *β* = *R*_0_*μ/*(1 − *p*_*a*_ + *r*_*β*_ *p*_*a*_) (See SI. Section S2.2 for the detailed derivation).

### Epidemic model for protesters

Transmission at protests can be modeled as interactions between susceptible protesters from a county and the total infectious protesters from all counties. The SEIR epidemic dynamics for protesters from each county *i* in state *s* is described according to

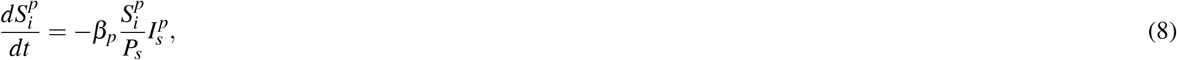

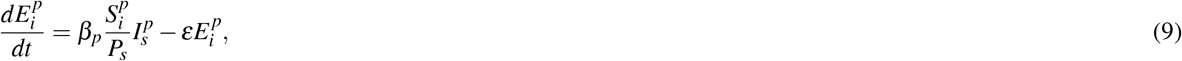

where 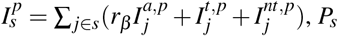 is the total number of protesters in *s*, and *β*_*p*_ is the transmission rate at protests. The superscript *p* denotes the compartments of protesters. The dynamics of the other compartments, 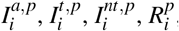, and 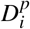, are the same with the dynamics in a simple SEIR model.

## Data Availability

All data produced in the present study are available upon reasonable request to the authors.

## Data Availability

All the original data can be downloaded from open repositories.

- **Protest**. The data is accessible from Count Love (https://countlove.org).
- **Mobility**. The commuting data can be downloaded from US Census (https://www.census.gov/data/tables/2015/demo/metro-micro/commuting-flows-2015.html), and the domestic aviation data can be obtained from the US Bureau of Transportation Statistics (https://transtats.bts.gov).
- **Intervention stringency**. The data is available from the Oxford Covid-19 Government Response Tracker (OxCGRT) (https://github.com/OxCGRT/USA-covid-policy).
- **COVID-19 statistics**. The COVID-19 statistics in the United States based on reports from state and local health agencies can be downloaded from The New York Times (https://github.com/nytimes/covid-19-data).

## Code Availability

The code for the full set of analysis may be available from the authors upon request.

## Akcnowledgements

M.C. was supported by the Ministry of Universities of the Government of Spain, under the program “Convocatoria de Ayudas para la recualificación del sistema universitario español para 2021-2023, de la Universidad Carlos III de Madrid, de 1 de Julio de 2021”.

## Author contributions

I.H., A.R. and M.C. conceived the study. I.H., A.R. and M.C. designed the study. I.H. developed the computational model. I.H. performed simulations. I.H. and L.F. analyzed data. I.H. wrote the draft. All authors edited the manuscript.

