## Supplementary Information for "Assessing the epidemic impact of protests during the COVID-19 pandemic"

#### S1. Data

**Protest data.** The protest data was retrieved from Count Love (https://countlove.org) on Jun. 29, 2021. The data covers protest events that occurred in the US between Jan. 1, 2017 and the present, based on crawling local newspaper and television sites on a daily basis [1]. For each protest event, the data has the information of date, location, attendees, tags, and source.

We analyzed the data since Jan. 1, 2020 as protests during the COVID-19 pandemic are of our interest. The resolution of "Locations" ranges from a specific place to a city, yet it is not standardized. Thus we used the location information at the state level in our analysis. "Attendees" reports the number of estimated participants in each protest.

"Tags" describes the purpose of a protest. Each protest event may have multiple tags. For the period later than Jan. 1, 2020, there are 614 unique tags in total, and 97 unique tags in the subset of protest events that contain "Coronavirus" in their tags. For these 97 unique tags, we manually identified the tags related to protests over COVID-19 policies. Those tags contain "Against pandemic intervention", "Against closure/relocation", and "Against vaccines". By doing so, we identified 783 protest events in 2021. For the period between Apr. 1, 2020 and Jun. 30, 2020, when the protests over COVID-19 were most active, there were 389 protest events. In the same period between Apr. 1, 2020 and Jun. 30, 2020, there were 4,121 protests with 1.66 million protesters in total including all types of protests.

**County-wise mobility.** The epidemic model with spatial spreading requires information of mobility between areas [2]. For this, we combined the county-wise commute data and the airport-wise aviation data to include both short- and long-distance travels. The county-wise commute data was obtained from 2011-2015 5-Year ACS Commuting Flows of US Census (https://www.census.gov/data/tables/2015/demo/metro-micro/commuting-flows-2015.html). We denote this commuting rate as $T_{ij}^{com}$.

The domestic aviation data was obtained from the US Bureau of Transportation Statistics for the travels in 2019 (https://transtats.bts.gov). As the aviation data is based on the number of travelers between US airports, it needs to be transformed into the county-wise mobility data. For this, we applied the gravity model on the aviation data, and reconstructed the county-wise travels based on the gravity model estimation. More precisely, the number of passengers $T_{ab}$ between a pair of airport $a$ and $b$ with distance $r_{ab}$ is estimated as $T_{ab} = GT_a^{\alpha}T_b^{\beta}/r_{ab}^{\gamma}$, where $T_a$ is the total outgoing traffic from airport $a$, $T_b$ is the total incoming traffic to airport $b$, and $\alpha$, $\beta$, $\gamma$ are the estimated exponents. As a result, the gravity model exponents are estimated as $\alpha = 0.4409$, $\beta = 0.4711$, and $\gamma = 0.2492$. Based on this observation, we reconstructed the county-wise aviation flows $T_{ij}^{air}$ using the gravity model on the population sizes $N_i$ and $N_j$ of counties $i$ and $j$, and their distance $r_{ij}$ according to

$$T_{ij}^{air} = T\frac{N_i^{\alpha}N_j^{\beta}}{r_{ij}^{\gamma}}, \tag{S1}$$

where $T$ is the normalization factor to satisfy $\sum_{i,j} T_{ij}^{air} = \sum_{a,b} T_{ab}$.

**Intervention stringency.** Since the COVID-19 pandemic, national and regional governments implemented many different countermeasures to mitigate the pandemic. The Oxford COVID-19 Government Response Tracker (OxCGRT) reports the stringency of these interventions as a time series data for the different levels of administration (https://github.com/OxCGRT/USA-covid-policy). We obtained the intervention stringency index data for US states [3] to study its relationship with protests over COVID-19 policies. This stringency index is based on the number of different countermeasures in effect. In this study, we used the index "StringencyIndexForDisplay" in the dataset as daily variations were removed in this index by moving average. See the original source for more details.

**COVID-19 and demographics.** The COVID-19 data in the United States based on reports from state and local health agencies was downloaded from The New York Times (https://github.com/nytimes/covid-19-data). The data contains the number of cumulative confirmed cases by counties. The demographic information of counties was obtained from the US Census Bureau (https://data.census.gov/cedsci/). Our simulation is based on the total 3141 counties excluding territories and two counties without commuting data.

### S2. Model

To model epidemic dynamics by protests, we need two different epidemic models, one for non-protesters, and the other for protesters. In this study, we used the SEIR (Susceptible-Exposed-Infectious-Recovered) compartmental disease transmission model [2] since it is one of the most simplest and widely-used models. As it is impossible to identify the home location of each protester with the dataset we have, we estimate the number of protesters in each county based on the protest data at the state level. Then, we simulate epidemic spreading among non-protesters and protesters separately, and aggregate the result. This loop is repeated for each day. Therefore, the simulation has a spatial resolution of a county, and a temporal resolution of a day.

To summarize, our coupled disease transmission model consists of four parts: 1) dividing the groups of non-protesters and protesters, 2) modeling epidemics for non-protesters, 3) modeling epidemics for protesters, and 4) aggregation. The steps 1) to 4) are repeated for each day. The details are presented below.

#### S2.1. Protesters and non-protesters

The protest data gives information of the number of total protesters at the state level each day, but the information of home counties of protesters is required for our county-level epidemic model. We estimate the home counties of protesters using the state-level protest data and the population distribution. We assume that the protesters in each state gather at a site in the state where their home counties are located, and the distribution of the home counties follows the population distribution of the home counties. More specifically, the number of protesters $P_i(t)$ on day $t$ from county $i$ in state $s$ is assumed to be

$$P_i(t) = P_s(t)\frac{N_i(t)}{\sum_{i\in s} N_i(t)}, \tag{S2}$$

where $P_s(t)$ is the total protesters in state $s$ on day $t$ known from the protest data, and $N_i(t)$ is the population of county $i$. Accordingly, the number of non-protesters is given as $Q_i(t) = N_i(t) - P_i(t)$.

#### S2.2. Baseline epidemic model for non-protesters

**Spatial SEIR epidemic model.** For modeling disease transmission among non-protesters, we used the SEIR epidemic model with domestic mobility. The theoretical design of this model is based on the global epidemic and mobility (GLEaM) model [2], except that we only considered domestic mobility instead of global mobility. In the SEIR epidemic model, the population is divided into the compartments of susceptible ($S$), exposed ($E$), infectious ($I$), recovered ($R$) and dead ($D$) populations. To incorporate spatial spreading between different areas, the infectious population is divided into three subgroups [2]: the asymptomatic infectious ($I^a$) population, the symptomatic population with traveling ($I^t$), and the symptomatic population without traveling ($I^{nt}$). The fractions of these subgroups are controlled by parameters $p_a$ and $p_t$ as $I^a = p_a I$, $I^t = (1-p_a)p_t I$, and $I^{nt} = (1-p_a)(1-p_t)I$. We set $p_a = 0.2$ and $p_t = 0.5$, and the transmission reduction factor of asymptomatic individuals $r_\beta = 0.5$ according to the references [2, 4].

To model spatial spreading, we also need a dataset of travels between counties. For this, we combine the county-wise commute data and the airport-wise aviation data to include both short and long travels (see S1 for the details). These travels can be decreased by mobility restrictions, mostly implemented in March, 2020. In our simulation, we assume that the mobility flow in our scope is 60% of the pre-COVID mobility, i.e., $r_m = 0.6$, based on the report on mobility reduction [5].

From the county-wise daily travel matrix $T_{ij}$, the visiting rate from county $i$ to county $j$ is defined as $\sigma_{ij} = T_{ij}/N_i$, where $N_i$ is the population of $i$ [2]. When we consider the returning rate $\tau$ from visiting, the effective number of individuals visiting from $i$ to $j$ is given as $\sigma_{ij}/\tau$. The returning rate $\tau$ can be different for commutes and airline travels since long-distance trips for business and vacation may have longer duration until travelers return to their home. Accordingly, we assume the returning rate of commutes as $\tau_{com} = 3$ day$^{-1}$ and $\tau_{air} = 1/14$ day$^{-1}$.

Therefore, the visiting rate $\sigma_{ij}$ and the effective number of visitors $v_{ij}$ are given as

$$\sigma_{ij} = \frac{T^{com}_{ij} + T^{air}_{ij}}{N_i}, \tag{S3}$$

$$v_{ij} = \frac{1}{N_i}\left(\frac{T^{com}_{ij}}{\tau_{com}} + \frac{T^{air}_{ij}}{\tau_{air}}\right). \tag{S4}$$

In addition, we denote the summation of $v_{ij}$ over all origins as $v_j = \sum_i v_{ij}$.

Here we summarize the mathematical formula needed for our computational model, which was first introduced by Balcan *et al* [2]. For each county $i$, the rate of change of each compartment is given as

$$\frac{dS_i}{dt} = -\lambda_i S_i, \tag{S5}$$

$$\frac{dE_i}{dt} = \lambda_i S_i - \epsilon E_i, \tag{S6}$$

$$\frac{dI^a_i}{dt} = \epsilon p_a E_i - \mu I^a_i - \nu I^a_i, \tag{S7}$$

$$\frac{dI^t_i}{dt} = \epsilon(1-p_a)p_t E_i - \mu I^t_i - \nu I^t_i, \tag{S8}$$

$$\frac{dI^{nt}_i}{dt} = \epsilon(1-p_a)(1-p_t)E_i - \mu I^{nt}_i - \nu I^{nt}_i, \tag{S9}$$

$$\frac{dR_i}{dt} = \mu(I^a_i + I^t_i + I^{nt}_i), \tag{S10}$$

$$\frac{dD_i}{dt} = \nu(I^a_i + I^t_i + I^{nt}_i). \tag{S11}$$

Here, the transmission rate $\lambda_j$ given in the reference [2] can be simplified as

$$\lambda_j = \frac{\beta_j}{(1+\sigma_j/\tau)N^*_j}\left[I^{nt}_j + I^e_j\right] + \frac{1}{(1+\sigma_j/\tau)}\sum_{i\in\nu(j)}\left[\frac{\beta_j\sigma_{ij}}{N^*_j\tau}I^e_i + \frac{\beta_i\sigma_{ji}}{N^*_i\tau}\left(I^{nt}_i + I^e_i + \sum_{l\in\nu(i)}\frac{\sigma_{li}}{\tau}I^e_l\right)\right], \tag{S12}$$

$$= \frac{\beta_j}{(1+v_j)N^*_j}\left[I^{nt}_j + I^e_j\right] + \frac{1}{(1+v_j)}\sum_{i\in\nu(j)}\left[\frac{\beta_j}{N^*_j}v_{ij}I^e_i + \frac{\beta_i}{N^*_i}v_{ji}\left(I^{nt}_i + I^e_i + \sum_{l\in\nu(i)} v_{li}I^e_l\right)\right], \tag{S13}$$

where $I^e_i = \frac{I^t_i + r_\beta I^a_i}{1+v_i}$ and $N^*_j = I^{nt}_j + \frac{N_j - I^{nt}_j}{1+v_j} + \sum_{i\in\nu(j)}\frac{N_i - I^{nt}_i}{1+v_i}v_{ij}$. We assume that the transmission rate without mobility $\beta_j$ is uniform across different counties. Then, the transmission rate can be estimated from the reproduction number $R_0$ as $\beta = R_0\mu/(1-p_a+r_\beta p_a)$.

**Estimation of the initial epidemic size.** The SEIR model needs an initial epidemic size as epidemic spreading is driven by interactions between the susceptible population and the infectious population. In this study, we focused on the epidemic dynamics between Apr. 1, 2020 and Jun. 30, 2020 since protests were most active in this period. Therefore, we used the estimated epidemic size on Apr. 1, 2020 as the initial epidemic size for the model.

Since the data on the epidemic trend only contains the number of cumulative infected cases by testing, the number of exposed individuals needs to be estimated. Let the number of cumulative infected cases on day $t$ in county $i$ from the data be $C_i(t)$. Then, its daily difference $I^{new}(t) = \Delta C_i(t)/\Delta t$ is equal to the incoming flux from the exposed cases $\epsilon E_i$ by definition in Eq. (S7). Due to the weekly periodicity by the small number of testing in weekends, we calculated $I^{new}(t)$ based on its moving averages for previous 7 days. Then, we estimated $E^{est}_i(t)$ using the average daily increase in the next $\Delta t$ days to suppress the daily variations as

$$E^{est}_i(t) = \frac{1}{\epsilon}\frac{\sum_t^{t+\Delta t} I^{new}(t')}{\Delta t}. \tag{S14}$$

We also set $\Delta t$ as 7 days considering the seasonality by weeks. The number of infectious individuals can be estimated using a non-spatial SEIR model. In the non-spatial SEIR model, the rate equation of exposed individuals

| Parameter | notation | value |
|---|---|---|
| base reproduction number (non-protesters) | $R_0$ | 1.05 |
| progression rate | $\epsilon$ | 1/5 |
| recovery rate | $\mu$ | 1/20 |
| death rate | $\nu$ | 0.0004 |
| proportion of asymptomatic patients | $p_a$ | 0.2 |
| proportion of traveling symptomatic patients | $p_t$ | 0.5 |
| relative infectiousness of the asymptomatic | $r_\beta$ | 0.5 |
| returning rate of commutes | $\tau_{com}$ | 3 |
| returning rate of air travels | $\tau_{air}$ | 1/14 |
| mobility reduction by lockdowns | $r_m$ | 0.6 |

**Supplementary Table S1:** Epidemic parameters for simulations.

is given as $dE/dt = \beta SI/N - \epsilon E$, similar to Eq. (S6). At the early beginning of disease spreading, we may assume that most population is susceptible as $S \approx N$. Then, using the estimated size of exposed population $E_i^{est}$ of each county, we get

$$I_i^{est}(t) \approx \frac{1}{\beta}\left(\frac{dE_i^{est}}{dt} + \epsilon E_i^{est}(t)\right), \tag{S15}$$

$$= \frac{1}{\beta}\left(\frac{E_i^{est}(t+\Delta t) - E_i^{est}(t)}{\Delta t} + \epsilon E_i^{est}(t)\right), \tag{S16}$$

where $\Delta t$ was also set to 7 days to suppress daily variations. The size of the susceptible population can be calculated as $S_i^{est}(t) = N_i - E_i^{est}(t) - I_i^{est}(\mathrm{t})$. In addition, the number of asymptomatic, traveling symptomatic, and non-traveling symptomatic individuals are given by their fractions as $I_i^a(t) = p_a I_i^{est}(t)$, $I_i^t(t) = (1-p_a)p_t I_i^{est}(t)$, and $I_i^{nt}(t) = (1-p_a)(1-p_t)I_i^{est}(t)$. Then, the epidemic dynamics is simulated using these initial compartment sizes for Apr. 1, 2020.

**Baseline epidemic dynamics.** This study used the SEIR epidemic model with mobility to simulate the epidemic dynamics of non-protesters. This baseline epidemic model without protests needs to be adjusted to describe the epidemic dynamics in the data.

Based on the initial epidemic size on Apr. 1, we simulated the epidemic dynamics by changing the base reproduction number by a factor of 0.05 to reconstruct the dynamics between Apr. 1, 2020 and Jun. 30, 2020, when the protests were most active. As a result, the reproduction number of 1.05 best described the dynamics observed in the data for the period before the surge in June, 2020. Supplementary Figure S1 shows the baseline epidemic dynamics simulated by our model without protests for different reproduction numbers.

To simulate the epidemic dynamics among non-protesters and protesters separately, we need to estimate the size of each epidemic compartments of non-protesters and protesters. We assume that the proportion of each compartmental population is the same for non-protesters and protesters. As the number of protesters and non-protesters on day $t$ in county $i$ are given as $P_i(t)$ and $Q_i(t)$ in Section S2.3, the compartment size can be estimated by their ratio. For each element in the set of compartments $X_i^m(t) \in \{S_i(t), E_i(t), I_i^a(t), I_i^t(t), I_i^{nt}(t)\}$, the compartment size $X_i^{m,np}$ of non-protesters is given as

$$X_i^{m,np}(t) = X_i^m(t)\frac{Q_i(t)}{Q_i(t) + P_i(t)}. \tag{S17}$$

In the same way, the compartment size $X_i^{m,p}$ of protesters is given as

$$X_i^{m,p}(t) = X_i^m(t)\frac{P_i(t)}{Q_i(t) + P_i(t)}. \tag{S18}$$

#### S2.3. Epidemic model for protesters

**Protest-SEIR model.** Our model supposes that protesters from their home counties gather at an arbitrary site in the state. In this case, the epidemic dynamics can be simulated with a simple non-spatial SEIR epidemic model. During a protest, protesters from each county can interact with infectious protesters, and all protesters return to their home counties after the protest. Thus, transmission of the disease can be modeled as interactions between

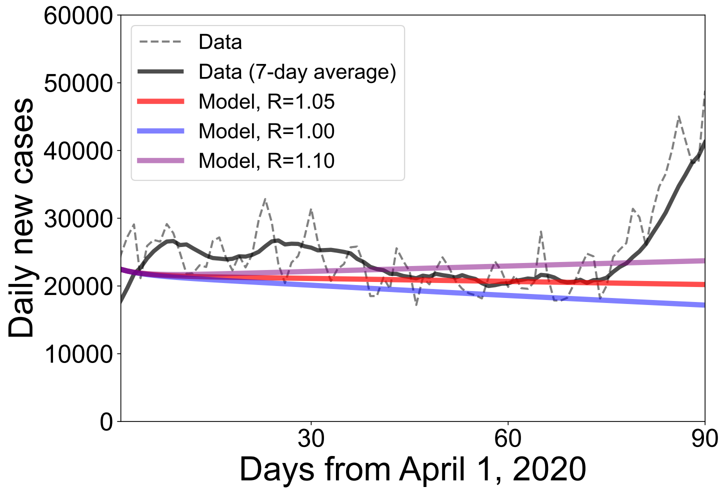


**Supplementary Figure S1:** Baseline epidemic model for non-protesters. The black curve shows the total daily new cases over the entire US for 90 days from April 1, 2020 (dotted for the raw data and solid for 7-day averages). The red, blue and purple curves describe the trend simulated by our epidemic model for the reproduction number of 1.05, 1.00 and 1.10, respectively.

susceptible protesters $S_i^p$ from county $i$ and the total infectious protesters $I_s^p$ from all counties in the same state $s$. The following equations describe the SEIR epidemic dynamics for each county $i$ in state $s$ according to

$$\frac{dS_i^p}{dt} = -\beta_p \frac{S_i^p}{P_s} I_s^p, \tag{S19}$$

$$\frac{dE_i^p}{dt} = \beta_p \frac{S_i^p}{P_s} I_s^p - \epsilon E_i^p, \tag{S20}$$

where $I_s^p = \sum_{j \in s}(r_\beta I_j^{a,p} + I_j^{t,p} + I_j^{nt,p})$, $P_s$ is the total number of protesters in $s$, and $\beta_p$ is the transmission rate at protests. The superscript $p$ denotes the epidemic compartments of protesters. The dynamics of the other compartments, $I_i^{a,p}$, $I_i^{t,p}$, $I_i^{nt,p}$, $R_i^p$, and $D_i^p$, are the same with the dynamics in Eqs. (S7-S11). As input for this epidemic model, the size of each compartment is estimated by its proportion according to Eq. (S18).

**Transmission rate at protests.** To simulate the epidemic dynamics with the above model, we need the value of the transmission rate at protests, $\beta_p$. Since there is no empirical study about the transmission rate of COVID-19 at protests, the value should rely on plausible assumptions. For these assumptions, there are two important factors: 1) the outdoor transmission rate and 2) the crowd density.

There is no consensus about the outdoor transmission rate of COVID-19 due to the lack of studies, but a review reported that the odds of indoor transmission is 18.7 times (6 - 58 times, 95% confidence interval) higher than outdoor transmission [6]. According to this report, we consider that the outdoor transmission rate is 1/18.7 of the indoor (normal) transmission rate, and its upper and lower bounds are 1/6 and 1/58. This low outdoor transmission rate necessarily leads to less infections at street protests.

Another important factor is the crowd density. While low outdoor transmission can be a plus for epidemiological safety of protests, the high crowd density at protests can be a great risk. The high crowd density can be roughly estimated using the Jacobs Crowd Formula [7, 8]. According to the formula, a light crowd has one person per 10 square feet and a dense crowd has one person per 4.5 square feet. As the distance of 2m is usually considered as a safe distance for social distancing, we calculated the number of individuals within a radius of 2m. As a result, the number ranges from 13.5 to 30, which means that there are 12.5 - 29 (i.e., 20.75 on half) other people excluding the one at the center. As most of daily communications in a close distance are one-to-one conversations, we consider that a protester has 20.75 times more close contacts with the others, and its lower and upper bounds are 12.5 and 29.

Combining these two factors, i.e., the low outdoor transmission rate and the high crowd density, we estimate the reproduction number at protests, and its upper and lower bounds. As a result, the base reproduction number at protests is thought to be 1.11 times higher than the normal reproduction number, and its upper and lower bounds are 0.216 and 4.83 times. Since many of protests during the pandemic, e.g., protests over COVID-19 policies, are often contrary to social distancing measures, we may consider that government interventions are not much effective for protesters. The normal reproduction number without interventions at the beginning of the pandemic [9] was around 3, and therefore we get the base reproduction number at protests $R_p^o$ as 3.33 and its upper and lower bounds as $R_p^l = 0.65$ and $R_p^u = 14.5$. Like the transmission rate for non-protesters, the transmission rate at protests is obtained as $\beta_p = R_p\mu/(1 - p_a + r_\beta p_a)$. The epidemic model simulates the epidemic dynamics on these three reproduction numbers.

##### S2.4. Aggregation

For each day, the compartmental population of the SEIR model is divided into the groups of protesters and non-protesters by their proportions in the protest data according to Eqs. (S17) and (S18). Then, the epidemic models for non-protesters and protesters calculate the epidemic spreading separately, and the differences are summed up at the county level. These steps of division, calculation and aggregation are repeated for each day until the simulation ends.

#### S3. Scaling analysis

##### S3.1. Multiplied epidemic trend

To estimate protest-driven infections at different epidemic situations, we multiplied the number of daily COVID incidence in the data for the period from April 1, 2020 to July 1, 2020, and adjusted the baseline reproduction number to describe this multiplied epidemic trend. The multiplication factor ranges from 1 to 15, which corresponds to daily incidence from about 20,000 to 300,000.

Since this multiplication changes the initial size of each compartment in the epidemic model, the reproduction number in the baseline epidemic model needs to be adjusted to properly describe the multiplied epidemic trend. The reproduction number that best describes each multiplied trend slightly increases with the increasing multiplication factor, ranging from 1.05 (for no multiplication) to 1.20 (for 15 times multiplication). Supplementary Figure S2 shows the multiplied epidemic trend and the model estimation for each multiplication factor.

##### S3.2. Simple scaling relation of protest-driven infections

Disease transmission in protests occurs between susceptible protesters and infected protesters with a reproduction number $R_p$ ranging from 0.65 to 14.5. Therefore, we can make a simple conjecture in which excess infected cases by protests are proportional to the protest size, baseline daily incidence, and $R_p - R_0$, where $R_0$ is the baseline reproduction number for non-protesters. This conjecture is consistent with the simulation result of new excess cases per day proportional to the protest size and the average daily cases (Fig. 3c in the main text).

Here we derive a simple scaling relation of protest-driven infections over the entire country based on the conjecture. For the total number of daily protesters $P$, the total population $N$, and the daily new incidence $\Delta I_t$, the excess daily incidence driven by protests $\Delta I_p$ can be estimated as:

$$\Delta I_p \sim (R_p - R_0)\left(\frac{P}{N}\right)^\alpha \Delta I_t^\beta, \tag{S21}$$

and equivalently,

$$\log\frac{\Delta I_p}{R_p - R_0} \sim \alpha \log\left(\frac{P}{N}\right) + \beta \log \Delta I_t, \tag{S22}$$

where $\alpha$ and $\beta$ are the coefficients for unknown non-linearity of the variables. The validity and the non-linearity of this model can be evaluated using a linear regression analysis.

The result of the regression analysis in Supplementary Table S2 confirms that the scaling relation is linear and that the conjecture is valid. Therefore, the scaling relation in Eq. (S21) can be rewritten as

$$\Delta I_p \simeq C(R_p - R_0)\left(\frac{P}{N}\right)\Delta I_t, \tag{S23}$$

where $C$ is a constant. In this relation, $C$ is the only unknown variable. From the simulation result of multiplied protests and epidemics in Fig. 3c in the main text, we can estimate the constant $C$ for the maximum daily infections. As a result, $C$ is estimated as 0.331 from a regression analysis on $\Delta I_p$ and $(R_p - R_0)\left(\frac{P}{N}\right)\Delta I_t$.

Supplementary Figure S3 confirms the linear scaling relation and the constant by comparing the excess daily cases estimated by the scaling relation in Eq. (S23) (i.e., x-axis) and the excess cases estimated by simulations (i.e., y-axis).

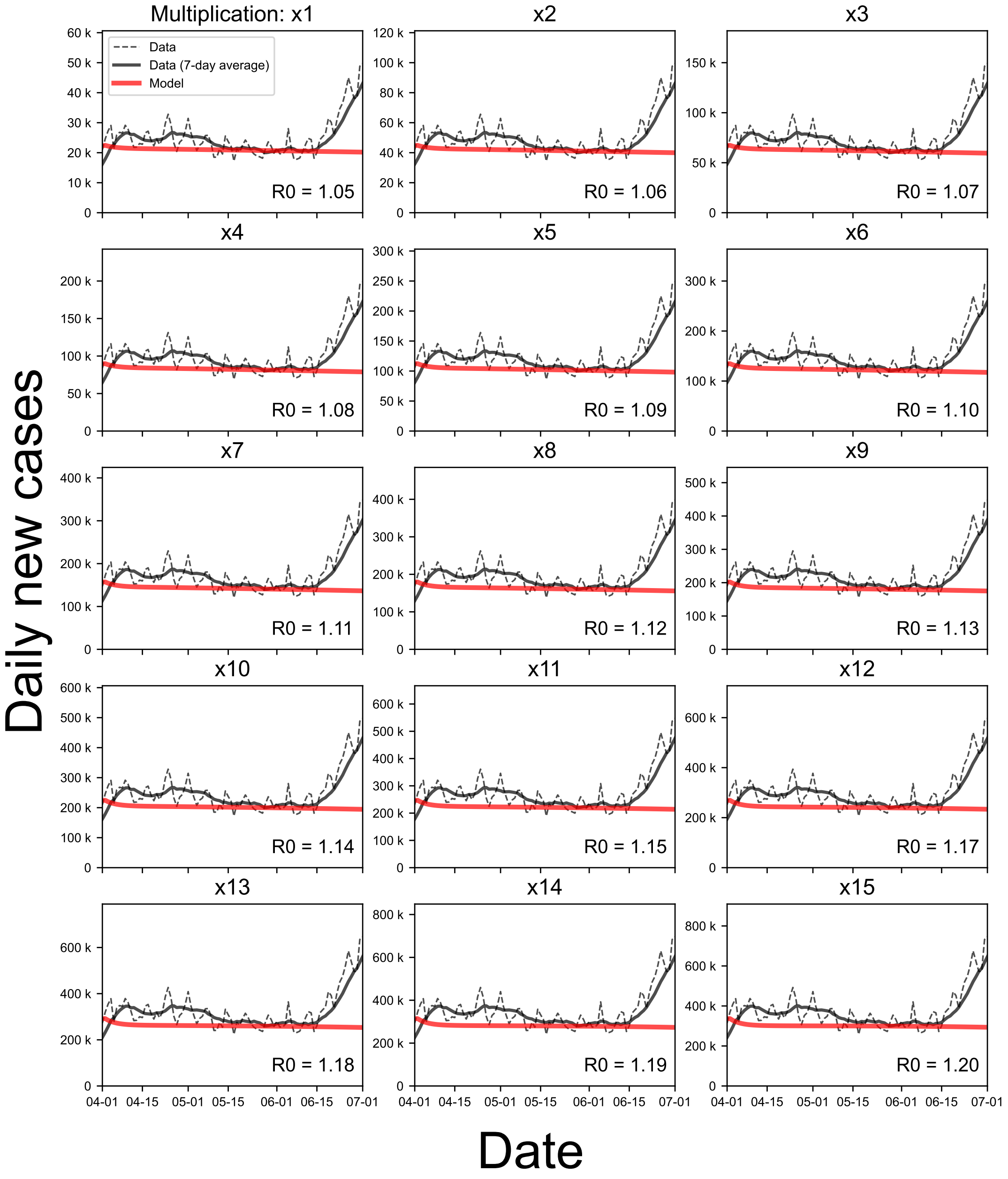


**Supplementary Figure S2:** Baseline epidemic model for multiplied epidemic trends. Each subplot shows the result of the baseline epidemic model for multiplied epidemic trends by different multiplication factors ranging from 1 to 15. The black curves show the total daily new cases over the entire US from April 1, 2020 to July 1, 2020 (dotted for the multiplied data and solid for the 7-day averages). The red curves describe the trend simulated by our epidemic model for the reproduction number that best describes the trend. The multiplication factor and the reproduction number are shown in each subplot.

#### S3.3. Critical protest size from the scaling relation

Equation (S23) can be used to calculate the precise critical protest size that induces an increase in daily incidence by a certain significant fraction to the total incidence in population. For the fraction $f = \Delta I_p/\Delta I_t$, we get,

$$P_c \simeq \frac{fN}{C(R_p - R_0)}. \tag{S24}$$

For $C = 0.331$ and a critical fraction of one percent, i.e., $f = 0.01$, we can calculate the critical size as a function of the reproduction number. Supplementary Figure S4 shows that the critical protest size in the US to make one percent increase in daily incidence may range from 0.745 million to 4.39 million in a total population of the US,

| Variables | coefficient | p-value |
|---|---|---|
| $\log P/N$ | 1.0019 | $< 0.001$ |
| | (0.003) | |
| $\log \Delta I_t$ | 1.0119 | $< 0.001$ |
| | (0.005) | |
| constant | -1.3198 | $< 0.001$ |
| | (0.065) | |
| Observations | 675 | |
| Adjusted $R^2$ | 0.996 | |

**Supplementary Table S2:** Regression analysis for the scaling relation of protest-driven epidemics. The dependent variable is $\log \frac{\Delta I_p}{R_p - R_0}$. Each value denotes the regression coefficient and the standard error is in parentheses.

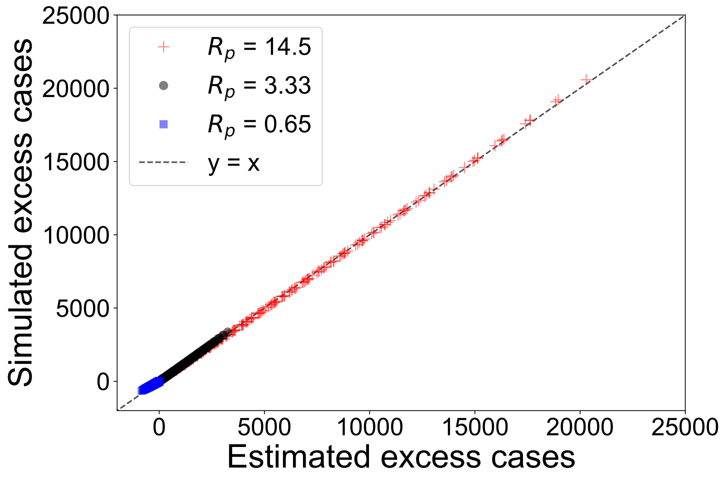


**Supplementary Figure S3:** Comparison of the analytical estimation and the simulation on the maximum daily protest-driven incidence. Each marker denotes a pair of a simulation result (i.e., y-axis) and the corresponding estimation (i.e., x-axis) by the scaling relation for a single parameter set of protest multiplication, epidemic multiplication and reproduction number (red for $R_p^u = 14.5$, black for $R_p = 3.33$ and blue for $R_p^l = 0.65$). The dotted black line is for $y = x$ that describes perfect matching of the scaling estimation and the simulation.

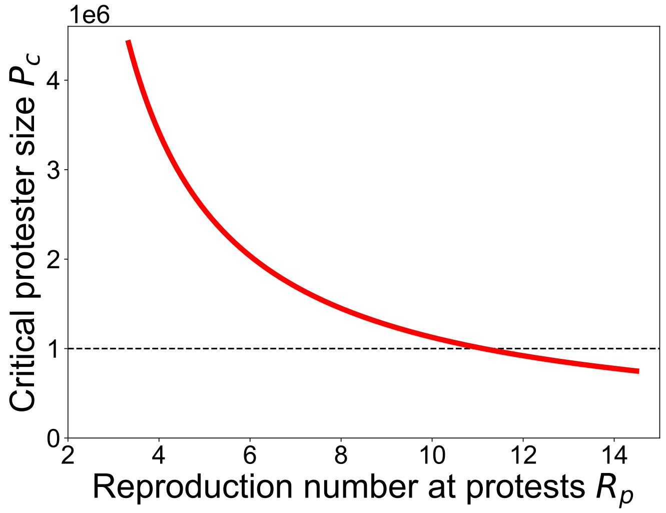


**Supplementary Figure S4:** Critical protest size as a function of the reproduction number at protests. The red line depicts the relation in Eq. (S24). The dotted black line denotes the critical size of 1 million.

i.e., 331 million, for varying reproduction numbers from $R_p^o = 3.33$ to $R_p^u = 14.5$. The reproduction number for the critical size of 1 million protesters is around $R_p = 11$.
